# Containing the Spread of Coronavirus Disease 2019 (COVID-19): Meteorological Factors and Control Strategies

**DOI:** 10.1101/2020.05.23.20111468

**Authors:** Jun Lin, Weihao Huang, Muchen Wen, Shuyi Ma, Jiawen Hua, Hang Hu, Dehong Li, Shan Yin, Yanjun Qian, Peiling Chen, Qiao Zhang, Ningbo Yuan, Shaolong Sun

## Abstract

The novel coronavirus disease 2019 (COVID-19) has spread globally and the meteorological factors vary greatly across the world. Understanding the effect of meteorological factors and control strategies on COVID-19 transmission is critical to contain the epidemic. Using individual-level data in mainland China, Hong Kong, and Singapore, and the number of confirmed cases in other regions, we explore the effect of temperature, relative humidity, and control measures on the spread of COVID-19. We found that high temperature mitigates the transmission of the disease. High relative humidity promotes COVID-19 transmission when temperature is low, but tends to reduce transmission when temperature is high. Implementing classical control measures can dramatically slow the spread of the disease. However, due to the occurrence of pre-symptomatic infections, the effect of the measures to shorten onset-to-isolation time is markedly reduced and the importance of contact tracing and quarantine and social distancing increases. The analytic results also highlight the importance of early intervention to contain the spread of COVID-19.

## Introduction

An outbreak of viral pneumonia caused by a novel coronavirus SARS-CoV-2 occurred in Wuhan, China in December 2019^1,2^. The novel coronavirus pneumonia (COVID-19) has exhibited high human-to-human transmissibility and rapidly spread to other Chinese provinces, as well as 213 other countries, areas, or territories^3,4^. As of 30 April 2020, a total of 3,090,445 confirmed cases and 217,769 deaths have been reported by WHO^4^. Wilson et al.^5^ estimated that different levels of control (via contact reduction) will cause 22% to 63% of the population in European countries sick, 0.20% to 0.60% hospitalized, and 0.07% to 0.28% dead.

At present, no effective treatment or vaccine is available for COVID-19. The only approaches to contain the spread of the epidemic are classical control measures, such as social distancing, isolation of confirmed cases, and contact tracing and quarantine. Understanding the effect of these control measures on COVID-19 transmission is critical for containing the spread of the virus. Moreover, the effectiveness of these control measures on virus transmission may vary with meteorological conditions such as temperature and relative humidity^6-8^.

To investigate the effect of meteorological factors and control measures on the spread of COVID-19, we first browsed and downloaded all reports published on the official websites of local health commissions in mainland China, Hong Kong, and Singapore. Then, we built a dataset containing the number of onset cases at the beginning of the outbreak in China, the individual information of travel history, symptom onset, seeking care at a hospital or clinic, and isolation date for all patients in 11 provinces in mainland China (Jilin, Liaoning, Heilongjiang, Tianjin, Henan, Shandong, Shaanxi, Guangxi, Yunnan, Anhui, and Chongqing), the individual information of all the cases confirmed from 1 to 30 April 2020 in mainland China, and the individual information reported by 17 March 2020 in Hong Kong and Singapore^3,9–22^. The other provinces in mainland China only partially disclosed the epidemic information of the patients confirmed before 1 April 2020. For Hong Kong and Singapore, certain epidemic information, such as isolation date, is not publicly available after 17 March 2020. We also obtained the doubling time of confirmed cases in the countries with no less than 180 cases by 25 March 2020^23^.

Using an extended susceptible–exposed–infectious–recovered (SEIR) model and the above data, we first estimated the transmission rates and effective reproduction numbers in mainland China and Hong Kong. Then, using the transmission rates of the provinces in mainland China and the doubling time of confirmed cases in 69 countries outside China, together with the daily temperature and relative humidity obtained from World Weather Online^24^, we analyzed the role of temperature and relative humidity in COVID-19 transmission. Thereafter, the effect of control measures is analyzed based on the detailed information on the time of symptom onset and isolation, and the estimated epidemiological parameters, such as transmission rate and pre-symptomatic transmission period. Finally, we evaluate the effect of classical control measures on COVID-19 transmission in different regions when the weather in the northern hemisphere warms up.

## Results

### Role of meteorological factors in COVID-19 transmission

COVID-19 is caused by severe acute respiratory syndrome coronavirus 2 (SARS-CoV-2), which is the seventh coronavirus known to infect humans^25,26^. Previous studies have shown that meteorological factors, such as temperature and humidity, may influence the survival and spread of human coronaviruses 229E, SARS-CoV, and MERS-CoV^6-8^. However, the role of temperature and humidity in COVID-19 transmission has not yet been clearly established.

The transmission rate of the virus is mainly affected by meteorological factors and social distancing measures. Almost the same level of control measures was implemented across the provinces in mainland China^27^. That is, the effect of social distancing measures is naturally controlled in the data. Therefore, using multiple regression, we clearly see the effect of temperature and relative humidity on the transmission rate of COVID-19. The regression results (Table 1) show that temperature has a negative effect and humidity has a positive effect on the transmission rate of COVID-19. The transmission rates are initially estimated using the extended SEIR model by setting the incubation period to 6.24 days, which is estimated based on our data. To check the robustness of the results in Table 1, we further estimate the transmission rates using the mean incubation periods of 5.80 days^28^ and 5.50 days^29^. The results are qualitatively the same, as shown in Tables S1 and S2 in supplementary information.

**Table 1.** Relationship between meteorological factors and COVID-19 transmission. The dependent variable is the estimated transmission rates of the provinces in China, which is derived by setting the incubation period to 6.24 days (Two-tailed test).

|  | **Coefficients** | **Standard Error** | **95% CI** | **P-value** |
| --- | --- | --- | --- | --- |
| Intercept | 0.012 | 0.044 | (-0.090, 0.113) | 0.800 |
| Temperature | -0.002 | 0.001 | (-0.003, -0.000) | 0.041 |
| Relative humidity | 0.001 | 0.001 | (0.000, 0.003) | 0.047 |
| R Square |  | 0.745 |  |  |
| Significance F |  | 0.042 |  |  |
| Observations |  | 11 |  |  |

Previous research of the human coronaviruses 229E shows that high relative humidity has a positive effect on coronavirus transmission when temperature is low but a negative effect when temperate is high^30^. The results in Table 1 are derived using the data from China when temperature is low. To develop a more holistic picture, we further analyzed the effect of temperature and relative humidity on the doubling time of confirmed cases in 69 other countries with at least 180 confirmed cases by 25 March 2020.

As the early cases in other countries are often imported, analyzing the local transmission of COVID-19 using the reported cases in the very beginning is inappropriate. Most cases in 8 of the 69 countries are imported until 60 cases are confirmed^23^. Therefore, to investigate the local transmission of the virus, we analyze the doubling time of confirmed cases when at least 70 cases have been confirmed in the countries. Moreover, the two countries, Russia and Pakistan, in which most cases are imported until 180 cases are confirmed are excluded from the sample^23^. We first use the duration from the 80th to 160th confirmed cases to represent the doubling time. The regression results (Table 2) show that temperature negatively relates to COVID-19 transmission. The effect of relative humility is moderated by temperature. High humidity promotes COVID-19 transmission when temperature is low but tends to reduce transmission when temperature is high. We also performed linear regression analyses using the durations from 70th to 140th and 90th to 180th confirmed cases and derived similar results (Tables S3 and S4).

**Table 2.**
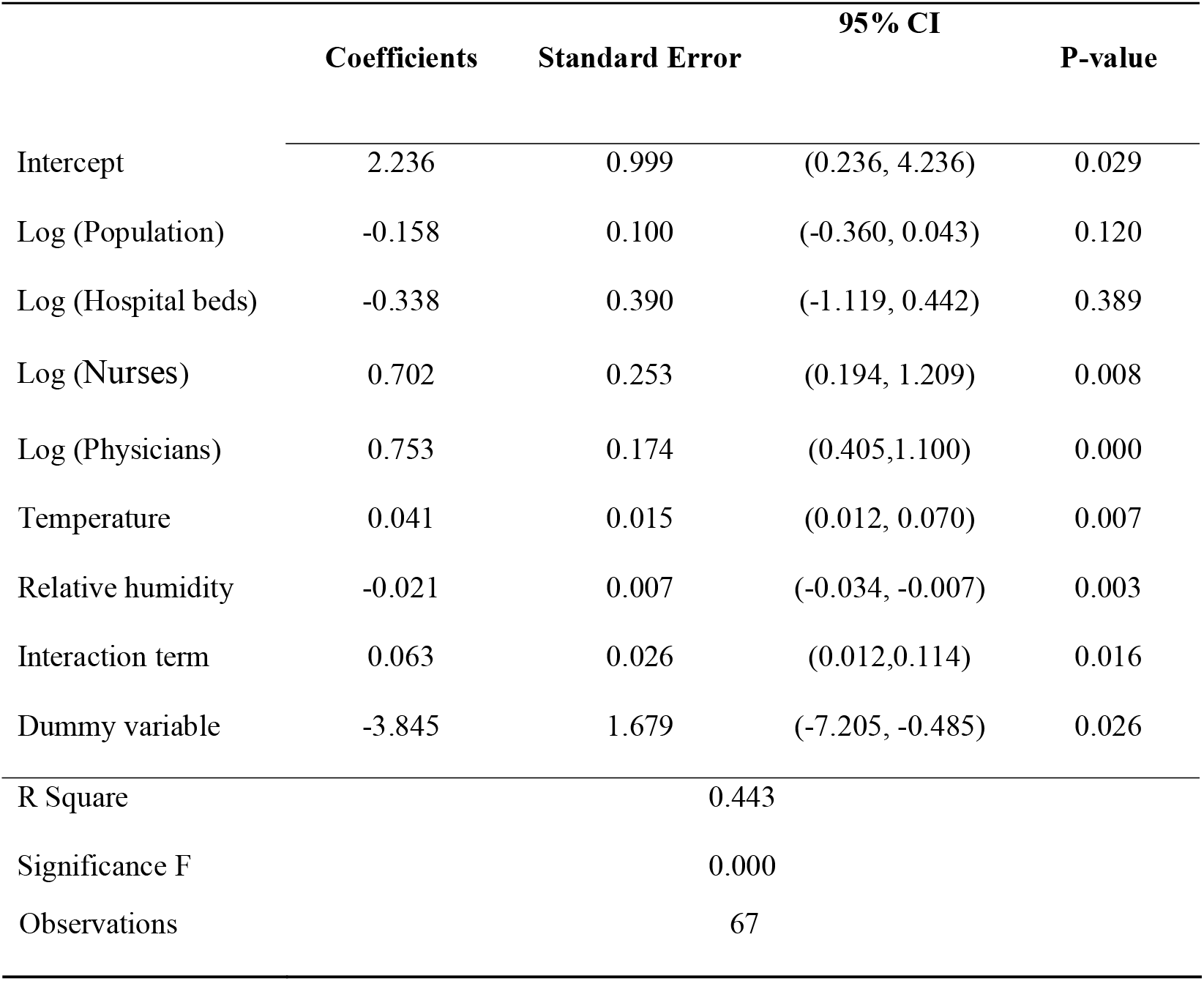
Relationship between meteorological factors and COVID-19 transmission. The dependent variable is the doubling time of confirmed cases (the duration from the 80^th^ to 160^th^ cases) in the countries outside China. The interaction term reflects the joint effect of high temperature and relative humidity (Two-tailed test).

### Measures to reduce the infectious period

Various control strategies can be applied to contain the spread of COVID-19. To find appropriate control strategies, we evaluate their effectiveness in reducing the effective reproduction number *R_e_*, which is defined as the average number of secondary cases per primary case. If *R_e_* exceeds one, the number of cases will increase over time and an outbreak of the disease is possible. If *R_e_* is less than one, then the disease will gradually die out. The effective reproduction number can be reduced in two ways: (1) by reducing the infectious period of infected individuals and (2) by reducing the person-to-person transmission rate. Rapid isolation of confirmed cases and timely contact tracing and quarantine are effective to reduce the infectious period, while social distancing measures, such as travel restrictions and contact precautions, help decrease the rate of disease transmission.

To examine the effect of classical control measures on infectious period reduction, we compared the average infectious period including the pre-symptomatic transmission time of the 11 provinces in mainland China before and after the first-level public health emergency response. The average infectious period was 9.42 days when few control measures were implemented. After most control measures were implemented, the average infectious period decreased to 4.20 days, representing a 55.42% decrease. Among all infected cases, 14.63% of them were quarantined before symptom onset, which contributed to 25.10% of total infectious period reduction. For the cases not quarantined, the infectious period was reduced to 4.84 days, accounting for 74.90% of total infectious period reduction.

However, such aggressive control measures to reduce the infectious period in mainland China may not be applicable to other regions because of the difference in cultural, political, and economic contexts. Therefore, we further examined the effectiveness of mild control measures such as those implemented in the early stage of the outbreak in Hong Kong and Singapore. We analyzed all the local cases until 29 February. After the mild control measures were implemented, the average infectious period in Hong Kong decreased from 12.76 to 8.13 days, representing a reduction of 36.30%. Only 4.76% of patients presenting symptoms were quarantined, which contributed to 11.88% of total infectious period reduction. The infectious period of cases not quarantined before the onset of symptoms was reduced to 8.48 days accounted for 88.12% of the total infectious period reduction. For Singapore, the results are similar. Only 1.64% of infected cases were quarantined before the onset of symptoms, and the infectious period decreased from 13.03 to 7.55 days after the control measures were implemented.

Notably, pre-symptomatic transmission plays a critical role especially when strict containment strategies are applied. When few control measures were taken in mainland China, 28.66% of secondary cases were infected by the primary cases not showing symptoms yet. However, after most control measures were implemented to control the epidemic, the percentage increased to 56.49%. This implies that the effective reproduction number is 129.85% higher than that without pre-symptomatic transmission. To a certain degree, this finding explains why COVID-19 is much more difficult to control than other coronavirus infections such as SARS and MERS, of which pre-symptomatic transmission rarely occurs.

Moreover, the detailed information on the 131 local cases presenting symptoms after 1 April 2020, when people in mainland China have returned to normal life, shows that because of pre-symptomatic transmission, the contribution of contact tracing and quarantine to the total infectious period reduction increases from 54.56% to 61.94%. On the contrary, the measures to shorten the onset-to-isolation time contributes 38.06% to the infectious period reduction compared with 45.44% when no pre-symptomatic transmission occurs. These results indicate that the effect of treatment and isolation measures for confirmed cases is markedly reduced and the importance of tracing and quarantine of close contacts increases because of pre-symptomatic infections.

### Measures to reduce transmission rates

Using the simulation results of mainland China, we evaluated the effectiveness of social distancing measures, such as compulsory mask wearing, closing most workplaces, and suspending public transport, in reducing the transmission rate. By fitting the number of onset cases of mainland China from 8 to 31 December, we found that the transmission rate in the early stage of the outbreak was 0.257, when the control measures were not implemented (Figure S1 and Table S5). Table S1 shows that the average transmission rate of the 11 provinces was 0.110, when most control measures were implemented. This implies that the implementation of strict control measures in mainland China reduced the transmission rate by 57.20%.

The relative effectiveness of social distancing increases because of pre-symptomatic infections. The reduction of *R_e_* is the combined effect of the reduction in transmission rate and infectious period. Social distancing measures could decrease *R_e_* by 57.20% whether pre-symptomatic infection occurs or not. However, the effectiveness of measures to reduce infectious period is weakened in the presence of pre-symptomatic infections. These measures could only decrease *R_e_* by 55.42% compared with 72.82% in the case of no pre-symptomatic transmission. Thus, the contribution of social distancing measures to contain the spread of COVID-19 becomes more prominent compared with the cases without pre-symptomatic infections.

### Timing of intervention

Early intervention is vital for containing the spread of epidemic. Using the epidemic and public health data of the 11 provinces in China, we investigated the effect of the number of infections before intervention on the infectious period of patients. The number of hospital beds per 10,000 people from China City Statistical Yearbook-2018 is used as a control variable. Logarithmic transformation of the variables was performed to normalize the distribution of values. The results show that the infectious period and thus the effective reproduction number *R_e_* increase with the number of infections before intervention (Table 3).

**Table 3.** The effect of the number of infections before intervention. The dependent variable is the average infectious period (days) of patients in each province (Two-tailed test).

|  | **Coefficients** | **Standard Error** | **95% CI** | **P-value** |
| --- | --- | --- | --- | --- |
| Intercept | 1.477 | 0.493 | (0.339, 2.615) | 0.017 |
| Log (Infections<br>before intervention) | 0.134 | 0.062 | (-0.010, 0.277) | 0.063 |
| Log (Hospital beds) | -0.669 | 0.306 | (-1.375, 0.036) | 0.060 |
| R Square |  | 0.506 |  |  |
| Significance F |  | 0.059 |  |  |
| Observations |  | 11 |  |  |

On the one hand, less medical resources can be used to treat newly infected cases if hospitals are already full of patients, which inevitably increases the onset-to-isolation time. On the other hand, large-scale community transmission becomes more possible as more people are infected before intervention, which increases the difficulty of tracing and quarantining contacts. Therefore, implementing control measures as early as possible is important to contain the spread of the disease.

## Discussion

We find that temperature negatively relates to COVID-19 transmission. However, the effect of relative humidity relies on temperature. High humidity promotes COVID-19 transmission when temperature is low but reduces COVID-19 transmission when temperature is high.

Currently, the temperature of most countries in the northern hemisphere is rising, which will gradually decrease the transmission of COVID-19. Specifically, regions such as Singapore and Philippines with high temperature and high humidity throughout the year have inherent advantages of containing the disease spread. In regions such as Japan, the temperature and humidity increase simultaneously. This greatly favors COVID-19 containment. In regions such as United States, England, and Spain, the relative humidity is expected to decrease when the weather becomes warmer. Although suppressing COVID-19 transmission in the regions is becoming more optimistic, the effect of changing meteorological conditions would be lower compared with the other regions in the northern hemisphere.

By contrast, the meteorological factors of countries in the southern hemisphere are becoming more unfavorable to contain the epidemic. For instance, in Australia and South Africa, control measures of the same intensity are expected to be less effective when the temperature decreases and the humidity increases. The meteorological data on the above countries for the past two years are shown in Figure 1.

**Fig. 1.**
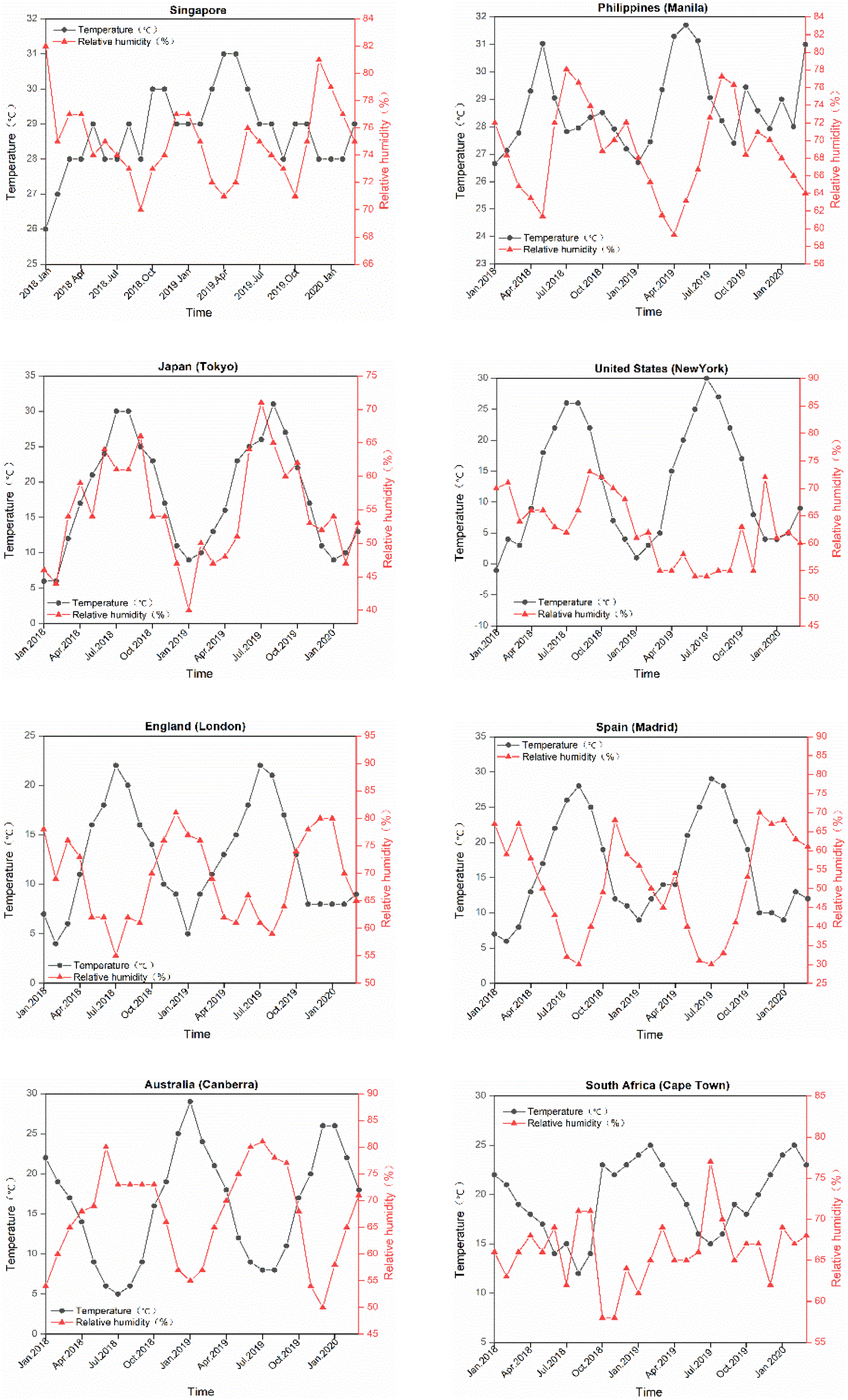
Temperature and relative humidity of eight representative countries.

The outbreak of COVID-19 in Singapore and Philippines shows that high temperature is not sufficient to contain the epidemic. Implementing stringent measures of social distancing, contact tracing and quarantine, and treatment and isolation of confirmed cases is important for controlling the COVID-19 transmission. Owing to the occurrence of pre-symptomatic infections, the effect of social distancing and contact tracing and quarantine is enhanced. In mainland China, these two types of measures contributed to 69.26% and 13.91% of the reduction in *R_e_*, respectively. Although placing a large number of people in quarantine, closing most workplaces, and suspending public transportation are costly, the epidemic cannot be controlled during a short period without these measures when there is already an outbreak of COVID-19. Both China and South Korea, the countries successfully controlled the spread of the diseases, implemented strict social distancing and contact tracing and quarantine measures, such as compulsory mask wearing and digital contact tracing.

However, if workplaces are not closed and individual movements are not restricted, then the transmission rate cannot be significantly reduced. In such cases, the epidemic still can be contained if measures to reduce the infectious period, such as contact tracing and quarantine, and treatment and isolation of confirmed cases, are strictly implemented. Most people in mainland China have returned to normal life since April 2020. Analyzing the local cases presenting symptoms after 1 April 2020 shows that the infectious period has been reduced to 2.57 days by quarantining 46.56% of the infected cases before symptom onset and shortening the onset-to-isolation time. The effective reproduction number is much lower than one in mainland China as the current transmission rate cannot be higher than 0.257, the transmission rate at the beginning of the outbreak. This result implies that if the control measures are well implemented, then the second round of COVID-19 outbreak in mainland China is almost impossible even when individual movements are not restricted. Analyzing the cases showing symptoms after 1 March 2020 in Hong Kong yields similar results. During this period, 38.46% of infected cases are quarantined before the onset of symptoms and the infectious period is reduced to 2.46 days. Moreover, through simulation, we find that the average transmission rate in Hong Kong is 0.074 (95% CI: 0.037–0.112) (Table S6). This implies that the outbreak of COVID-19 can be avoided by tracing and quarantining a large proportion of contacts and reducing the onset-to-isolation time.

This study has several limitations. First, our findings on the relationships between meteorological factors and the transmission rate of COVID-19 are derived from statistical and mathematical analyses using real-life data. Highly rigorous controlled medical experiments are still necessary to show their effect in a laboratory environment. Second, control measures are mainly evaluated in the context of mainland China, Hong Kong and Singapore. The differences in economics, politics, and even culture may lead to varying effects of control measures. Therefore, an issue that needs to be investigated in a global context is whether the control measures can be replicated outside these regions.

## Methods

### An extended SEIR model for estimating transmission parameters

We use an extended SEIR model to describe the transmission process of COVID-19. To the best of our knowledge, pre-symptomatic transmissions are possible for COVID-19, and a certain percentage of patients are quarantined before the onset of symptoms in some regions. Therefore, we extend the original SEIR model by adding the pre-symptomatic transmission process and the transmission process of the patients quarantined before symptom onset. We assume that a patient’s infectivity remains constant during the infectious period.

Figure 2 illustrates the transmission process. We divide the population into seven compartments. *S* is the susceptible population. *E* represents the patients who are in the incubation period but not infectious. The patients quarantined before the onset of symptoms flow from *E* into *I_ibo_* (transmission path 1), and the others flow into *I*_1−_*_iao_* (transmission path 2). *I_ibo_* and *I*_1__*_iao_* represent the patients who are pre-symptomatic but infectious. *R_ibo_* represents the patients who are isolated but not onset, and *I*_2_−*_iao_* represents the patients who are onset but not isolated. Finally, all the patients flow into *R*, representing those who are either isolated and symptom onset or recovered. The population dynamics are governed by the following system of differential equations:

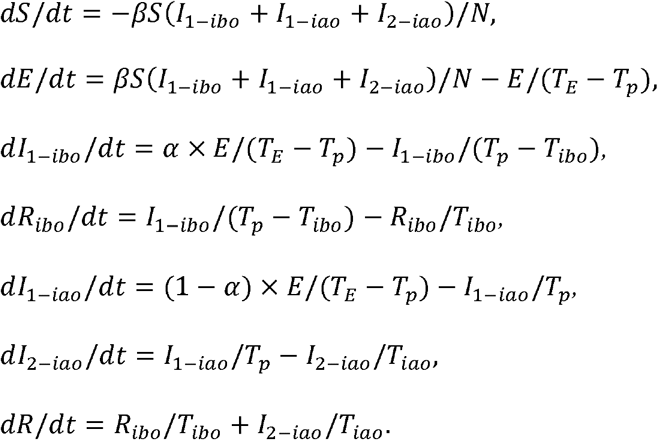

**Fig. 2.**
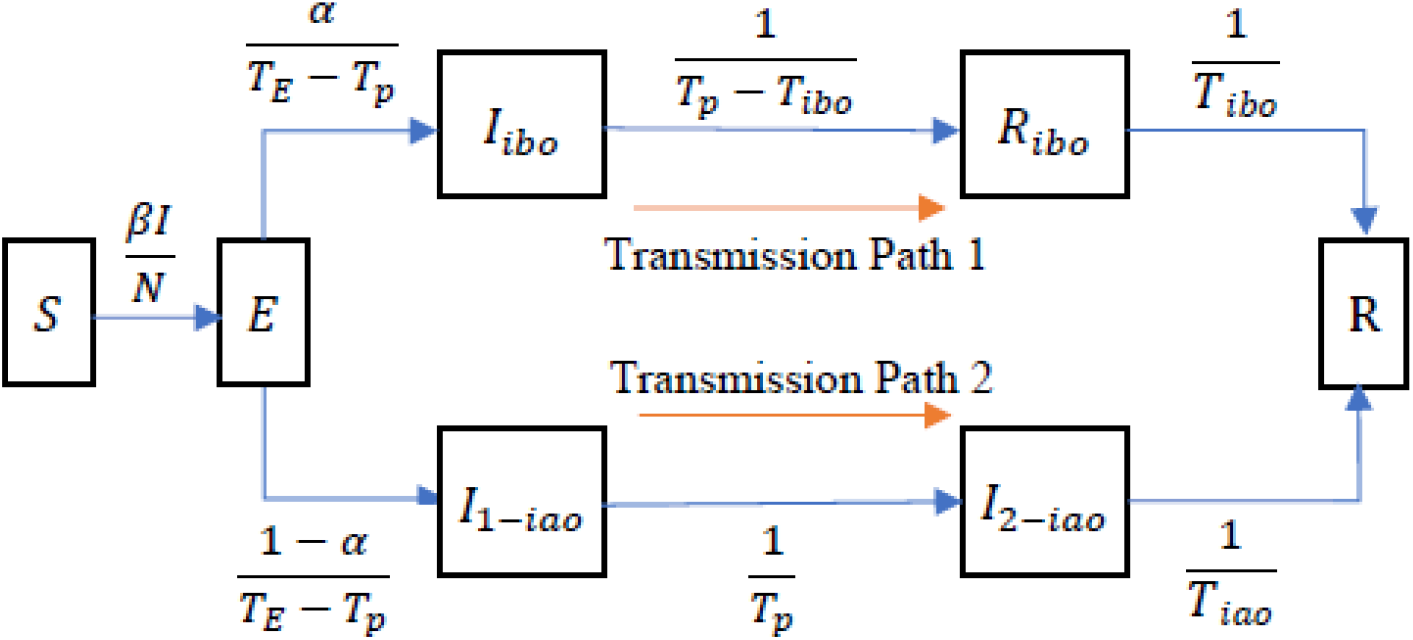
The transmission process of COVID-19.

The following parameters specify the flow rates. *N* is the population in the corresponding region; *β* is the transmission rate; *I* is the number of infectious cases, which is equal to *I_ibo_* + *I*_1_*_−iao_* + *I*_2_*_−iao_*; *T_E_* is the incubation period; *T_p_* is the pre-symptomatic transmission period; *α* is the rate of patients quarantined before the onset of symptoms; *T_ibo_* is the average isolated pre-symptomatic transmission time for the patients quarantined before the onset of symptoms; and *T_iao_* is the average onset-to-isolation time for the patients not quarantined. The values of input parameters, including *S*, *I*_1_*_−iao_*, *I*_2_*_−iao_*, *I_ibo_*, *R*, *T_E_*, *T_p_*, *α*, *T_ibo_*, and *T_iao_*, are extracted from the detailed information of the confirmed cases. The parameters *E*, *β*, and *T_p_*re estimated by the least squares method.

### Model Fitting

The model parameters best describing the onset data are identified using the least squares method. To avoid the influence of imported cases on local transmission, we only simulated the transmission process when the control measures were implemented and new imported cases were rare.

For the eleven provinces in mainland China, the time window of the simulation is from the first day after the first-level public health emergency response when the influence of imported cases on local transmission is negligible to 10 February 2020 after which few cases presenting symptoms. To understand the transmission process without intervention, we fitted the number of onset cases from 8 December 2019 when the first patient presenting symptom to 31 December 2019 when a cluster of COVID-19 disease cases caused by SARS-CoV-2 are first reported in mainland China^22^. For Hong Kong, we studied the local transmission of COVID-19 from 31 January 2020 when Hong Kong closed its port connecting mainland China to 6 March 2020 after which the number of onset cases may be underestimated due to delayed reporting. We used fifteen days’ onset data to fit the model; thus, the simulation period is from 31 January 2020 to 14 February 2020. Then, we started another simulation using the symptom onset data from 3 to 17 February 2020. Finally, we obtained eight values of transmission rate. Figures S1–S12 and Tables S6–S8 present the simulation results.

### Meteorological Factors

Data on temperature and relative humidity are collected from World Weather Online^24^. The meteorological data of the cities over the world are updated every three hours. The average of eight values (temperature or relative humidity) in one day is used as the daily value of a city.

The meteorological factors of the 11 provinces are used for the regression analysis on the transmission of COVID-19 in mainland China. To derive the meteorological factors of the provinces, we first counted the infected cases in the cities of the provinces before the first-level public health emergency response. The weight of a city is defined as the ratio of the infected cases in the city and the total infected cases in the corresponding province. The mean temperature (relative humidity) of a city are defined as the average temperature (relative humidity) during the time window of the simulation. For instance, the first-level public health emergency response was activated on 25 January 2020 in Shaanxi Province. Accordingly, the time window of the simulation is from 26 January 2020 to 10 February 2020, and the mean temperature and relative humidity are the average values during this period. Finally, the mean temperature (relative humidity) of a province is calculated as the sum of the products of the weights of the cities in the province and the corresponding average temperatures (relative humidities).

We also analyzed the effect of temperature and relative humidity on the epidemic doubling time in the countries with at least 180 confirmed cases by 25 March 2020^23^. To derive the meteorological factors of the countries, we first searched the initially reported cases in a country to determine the province where most of the early cases are infected. Then, we used the temperature and relative humidity of the provincial capital city to represent those of the country. To derive the temperature or relative humidity when the cases were infected, we need to estimate the median incubation period and the median time from symptom onset to laboratory confirmation. The average median incubation period based on our data and the relevant literature is five days ^27, 29, 31-33^. The average time from symptom onset to laboratory confirmation in mainland China, Hong Kong, and Singapore before implementing strict control measures is seven days. Therefore, we believe that most of the cases are infected around 12 days before the confirmation day. Specifically, the temperature and relative humidity during 12±3 days before the confirmation day of the cases are collected. Then the average temperature and relative humidity during the period are used in the regression analysis. For example, in Argentina, the 80^th^ case was confirmed on 19 March 2020 and the 160^th^ case was confirmed on 22 March 2020. Thus, the average temperature and relative humidity during 4 to 13 March in Argentina were calculated. Then, these data are used to analyze the effect of meteorological factors on the epidemic doubling time measured as the duration from the 80^th^ to 160^th^ confirmed cases.

### Effect of meteorological factors on COVID-19 transmission in other countries

We first use the transmission rates in eleven provinces in mainland China as the dependent variable in multiple regression. The intensity of local transmission and meteorological conditions in different provinces vary largely. Moreover, inter-provincial travel is restricted after the first-level public health emergency response, the highest state of emergency, is activated. Therefore, we can estimate the transmission rate in each province separately. We first extracted the key parameters of our SEIR model from the locally infected cases in each province, including the number of onset cases per day, incubation period, percentage of infected cases quarantined before the onset of symptoms, pre-symptomatic but infectious time of the cases quarantined before the onset of symptoms, and onset-to-isolation time of the cases not quarantined. Based on the extended SEIR model, the predicted onset case numbers and transmission rates of the provinces are derived as shown in Figures S2–S12, Table S5 and Tables S7-8. Then, we collected daily values of temperature and relative humidity during the simulation period for all the cities in the 11 provinces from World Weather Online^24^. The regression results are shown in Table 1.

Then, we use the doubling time of confirmed cases from the 80^th^ to 160^th^ confirmed cases in the countries outside China as the dependent variable in multiple regression. The population of a country and health resources such as the number of hospital beds, number of nurses, and number of physicians per 1,000 people are used as control variables. Logarithmic transformation of the doubling time and the control variables was performed to normalize the distribution of values. Temperature and relative humidity are the explanatory variables. We define a dummy variable as 1 if the temperature is above 25 ^8,30^; otherwise, 0. The interaction between the dummy variable and relative humidity is introduced to check whether the effect of humidity on disease transmission differs when the temperature becomes high. Table 2 presents the results. We also performed linear regression analyses using the durations from the 70^th^ to 140^th^ and 90^th^ to 180^th^ confirmed cases (Table S3–S4). Moreover, we used the temperature and relative humidity during 12±2 days before the confirmation days of the cases to further check the robustness of the findings. The results are qualitatively the same, as shown in Table S9.

### Infectious Period

For mainland China, the average infectious period of all patients with symptom onset before 20 January 2020 in the 11 provinces was used to indicate the infectious period before implementing control measures. This is because human-to-human transmission of COVID-19 was confirmed on 20 January 2020, and few control measures were implemented before this time. The first-level public health emergency response was activated on 24 or 25 January in the 11 provinces. Afterwards, the strict control measures were implemented. Given the lag between the time of implementing control measures and the time at which the measures take effect, we used the average infectious period of all patients with symptom onset between 1 and 10 February 2020 to indicate the infectious period after implementing control measures. Similarly, for Hong Kong and Singapore, local cases with symptom onset before 30 January 2020 and those between 5 and 29 February 2020 are used to estimate the infectious periods before and after implementing control measures, respectively.

The infectious period is composed of pre-symptomatic and symptomatic infectious periods. Through our simulation results, on average, the pre-symptomatic infectious is 2.70 days. If a patient is quarantined three days before the onset of symptoms, then the infectious period is 0. For the patients quarantined one and two days before the onset of symptoms, the infectious periods are 1.70 and 0.70 days, respectively. For cases who are not quarantined before symptom onset, their infectious period is the sum of the pre-symptomatic infectious period and the onset-to-isolation time.

## Data availability

All code and data are available in the supplementary materials and posted online at https://github.com/Ricardo-Wen/Containing-the-Spread-of-Coronavirus-Disease-2019-COVID-19-Meteorological-Factors-and-Control-Str.

## Acknowledgments

This work was supported in part by the National Natural Science Foundation of China (71672140 to J.L.), and in part by the National Social Science Foundation of China (16BGL017 to Y.Q.).

## Author contributions

J.L. developed the idea and research. W.H., M.W., S.M., J.H., H.H., D.L., S.Y. and N.Y. collected the epidemiological data. J.L., M.W. and P.C. conducted the analyses. W.H., D.L. and S.Y. wrote the first draft of the manuscript. J.L., Y.Q., Q.Z. and S.S. edited the manuscript. All authors read and approved the manuscript.

## References

1. Zhu, N. et al. A novel coronavirus from patients with pneumonia in China, 2019. N. Engl. J. Med. 382, 727–733 (2020).

2. Huang, C. et al. Clinical features of patients infected with 2019 novel coronavirus in Wuhan, China. Lancet 395, 497–506 (2020).

3. Li, Q. et al. Early transmission dynamics in Wuhan, China, of novel coronavirus–infected pneumonia. N. Engl. J. Med. 382, 1119–1207 (2020).

4. World Health Organization, Coronavirus Disease 2019 (COVID-19): Situation Report—101 (30 April 2020); https://www.who.int/docs/default-source/coronaviruse/situation-reports/20200430-sitrep-101-covid-19.pdf?sfvrsn=2ba4e093_2.

5. Wilson, N. et al. Modelling the Potential Health Impact of the COVID-19 Pandemic on a Hypothetical European Country. Preprint at https://www.medrxiv.org/content/10.1101/2020.03.20.20039776v1 (2020).

6. Van Doremalen, N., Bushmaker, T. & Munster, V. J. Stability of Middle East respiratory syndrome coronavirus (MERS-CoV) under different environmental conditions. Euro Surveill. 18, 20590 (2013).

7. Geller, C., Varbanov, M. & Duval, R. E. Human coronaviruses: insights into environmental resistance and its influence on the development of new antiseptic strategies. Viruses 4, 3044–3068 (2012).

8. Chan, K. H. et al. The effects of temperature and relative humidity on the viability of the SARS coronavirus. Adv Virol 2011, (2011).

9. The People’s Government of Jilin Province; http://www.jl.gov.cn/szfzt/jlzxd/.

10. Health Commission of Liaoning Province; http://wsjk.ln.gov.cn/.

11. Health Commission of Heilongjiang Province; http://feishizaixian.wt020.668895.com/.

12. Health Commission of Tianjin Municipal; http://wsjk.tj.gov.cn/.

13. Health Commission of Henan Province; http://hnwsjsw.gov.cn.

14. Health Commission of Shandong Province; http://wsjkw.shandong.gov.cn/.

15. Health Commission of Shandong Province; http://wsjkw.shandong.gov.cn/.

16. South China Morning Post; https://www.ngzb.com.cn/news/71324.html.

17. Health Commission of Yunnan Province; http://ynswsjkw.yn.gov.cn/wjwWebsite/web/index.

18. Health Commission of Anhui Province; http://wjw.ah.gov.cn/.

19. Health Commission of Chongqing Municipal; http://wsjkw.cq.gov.cn/.

20. The Government of the Hong Kong Special Administrative Region Press Releases; https://www.info.gov.hk/gia/general/202004/09.htm?fontSize=1.

21. Ministry of Health, Singapore; https://www.moh.gov.sg/covid-19/past-updates.

22. The Novel Coronavirus Pneumonia Emergency Response Epidemiology Team. The epidemiological characteristics of an outbreak of 2019 novel Coronavirus diseases (COVID-19) —China, 2020. China CDC Weekly 2, 113–122 (2020).

23. World Health Organization, Coronavirus disease (C0VID-2019) situation reports 1–65; https://www.who.int/emergencies/diseases/novel-coronavirus-2019/situation-reports/.

24. World Weather Online; https://www.worldweatheronline.com/.

25. Andersen, K.G., Rambaut, A., Lipkin, W. I., Holmes, E. C. & Garry. R. F. The proximal origin of SARS-CoV-2. Nat. Med. 26, 450–452 (2020).

26. Zhou, P. et al. A pneumonia outbreak associated with a new coronavirus of probable bat origin. Nature 579, 270–273 (2020).

27. Tian, H. et al. An investigation of transmission control measures during the first 50 days of the COVID-19 epidemic in China. Science 368, 638–642 (2020).

28. Backer, J. A., Klinkenberg, D. & Wallinga, J. Incubation period of 2019 novel coronavirus (2019-nCoV) infections among travellers from Wuhan, China, 20–28 January 2020. Euro Surveill. 25, 20–28 (2020).

29. Lauer, S. A. et al. The incubation period of 2019-nCoV from publicly reported confirmed cases: Estimation and application. Ann Intern Med. 172, 577–582 (2020).

30. Ijaz, M. K., Brunner, A. H., Sattar, S. A., Nair, R. C. & Johnson-Lussenburg, C. M. Survival characteristics of airborne human coronavirus 229E. J. Gen. Virol. 66, 2743–2748 (1985).

31. Pung, R. et al. Investigation of three clusters of COVID-19 in Singapore: implications for surveillance and response measures. Lancet 395, 1039–1046 (2020).

32. Guan, W. et al. Clinical Characteristics of Coronavirus Disease 2019 in China. N. Engl. J. Med. 382, 1708–1720 (2020).

33. Yang, Y. et al. Epidemiological and clinical features of the 2019 novel coronavirus outbreak in China. Preprint at https://www.medrxiv.org/content/10.1101/2020.02.10.20021675v2 (2020).

